# Association between Body Mass Index and Risk of Aortic Stenosis in Women

**DOI:** 10.1101/2023.09.26.23296191

**Authors:** Silvana Kontogeorgos, Annika Rosengren, Tatiana Zverkova Sandström, Michael Fu, Martin Lindgren, Carmen Basic, Maria-Teresia Svanvik, Demir Djekic, Erik Thunström

**Author notes:** **Corresponding author:** Silvana Kontogeorgos, PhD, Dept. of Molecular and Clinical Medicine/Cardiology, Sahlgrenska Academy, University of Gothenburg; Sahlgrenska University Hospital/Östra Hospital, Diagnosvägen 11, SE 416 50 Gothenburg, Sweden, Dept. of Clinical Physiology, Sahlgrenska University Hospital, Gothenburg, Sweden.; E-post.

## Abstract

**Background:** Overweight and obesity are increasing globally, as are life expectancy and disorders associated with aging, including calcific aortic stenosis (AS). Studies investigating the correlation between high body mass index (BMI) and AS are contradictory and inconclusive. This study examines a potential association between BMI and early AS in women.

**Methods:** By linking the Swedish Medical Birth Register (MBR) and the Swedish National Patient Register (NPR), we included women between 18 and 55 years with a first childbirth from 1982 to 2020. Diagnosis of subsequent AS and comorbidities were defined according to the International Classification of Diseases (ICD) 8, 9, and 10 codes. The women were divided into eight groups based on their BMI, with BMI 20 kg/m^2^-<22.5 kg/m^2^ as reference. Cox proportional hazards regression models were used to investigate the difference in the risk of being diagnosed with AS.

**Results:** The mean age at inclusion among 1,722,798 women was 28 years, and the mean BMI was 24 kg/m^2^. Some 21% were overweight (BMI 30 kg/m^2^-<35 kg/m^2^) and 10.7% were obese (BMI≥35 kg/m^2^). During a median follow-up of 19.5 years, 2,513 women (0.15%) were diagnosed with AS. The age-adjusted risk of being diagnosed with AS, compared to a reference group with BMI 20 -<22.5 kg/m^2^, increased with higher BMI to 2.8 (95%CI 2.43-3.24) times higher in women with BMI 30 -<35 kg/m^2^, and to 3.67 (95%CI 2.91-4.64) times higher in those with severe obesity (BMI ≥35 kg/m^2^). Similar results were found after excluding AS of rheumatic and congenital etiology.

**Conclusions:** Higher BMI correlated with a higher risk of developing AS in women, regardless of etiology, beginning with increased risk even within the upper normal range, reaching a three times higher risk in women with severe obesity.

**Clinical perspective:** What is new?

- This study investigates a subpopulation seldom included in valvular heart diseases studies, younger women, showing that women with overweight and obesity have approximately three times higher risk to be diagnosed with AS.
- Even women in the BMI group considered high-normal (BMI 22.5 -<25 kg/m^2^) had higher risk for AS compared those with BMI 20 -<22.5 kg/m^2^.

What are the clinical implications?

- The percentage of individuals with overweight and obesity increases and in the same time, AS becomes more prevalent as the population ages. Considering that there is no medical treatment that can ameliorate AS prognosis, prevention by maintaining a low-normal or normal BMI becomes even more important.

## Introduction

Degenerative aortic stenosis (AS) increases in prevalence rate with age ^1^ (and is expected to rise) ^2-4^. Among valvular heart diseases, AS is the single valvular heart disease leading to most interventions. Five-year mortality has been estimated at 56% in moderate and 67% in severe AS ^5^. No medical treatment improves AS prognosis or delays progression ^6-8^ and only aortic valve intervention improves survival.

Obesity rates are increasing ^9^, leading to major health problems. In Sweden, 40% of adult women are currently overweight (body mass index [BMI] between 25 and 30 kg/m^2^), and 10% are obese (BMI <30 kg/m^2^) ^10^. Obesity is associated with or predisposes to other health problems, including metabolic and cardiovascular, negatively affecting quality of life and life expectancy. A link between obesity and AS has been shown ^11^. In addition, in previous work from our group, we found that middle-aged men with obesity were predisposed to be diagnosed with AS later in life ^12,13^. However, although these studies had an extended follow-up, only men who were middle-aged at baseline were included. In addition, no large studies investigated whether the relationship between early life BMI and incident AS applies to the female population.

The correlation between higher BMI and AS has been examined in several studies, sometimes with contradictory results: some found a correlation between AS and BMI, whereas others did not. Some previous studies focused on the AS progression ^14-16^, others on aortic valve calcification ^17-19^ and others on AS ^11,20,21^.

These studies concluded that higher BMI correlated with faster AS progression rates, aortic valve calcification, and AS prevalence. However, several of these studies were limited by a cross-sectional design or risk of bias, including only older patients with severe AS. Other studies did not find a link between high BMI and AS ^22,23^ or, in a study on older adults, even found an inverse relationship ^24^. Because AS is a condition that commonly evolves over a long period, it is crucial to investigate to what extent an elevated body weight in young people predicts AS later in life. Therefore, to clarify this issue we included a nationwide population of younger women who had their weight recorded in early pregnancy to test the hypothesis that higher BMI is associated with a subsequent AS diagnosis in middle age.

## Methods

### Study population

From the Swedish Medical Birth Register (MBR), after exclusion according to the criteria described in Supplementary Figure 1, we identified all women aged 18-55 years with first registered childbirth between 1982 and 31 December 2020, regardless of whether it was the first pregnancy or not. The inclusion date was the childbirth date or, if missing, the discharge date minus 3 days or, if the latter was also missing, the date for the first antenatal visit (median 10 weeks of pregnancy) plus 207 days (the difference between the average duration of pregnancy in the population and 70 days) was used.

The nationwide Swedish MBR started in 1973 and records births in Sweden with a coverage of 99% ^25^. Between 1990 and 1991, no data about height and weight were recorded and women giving birth during those years were not included in the study. The study was approved by the Regional Ethics Review Board in Gothenburg and complies with the principles outlined in the Declaration of Helsinki.

### Variables: outcome, exposure, and covariates

The outcome in our study was incident AS defined as ICD codes (irrespective of position), ICD 8 (until 1986), ICD 9 (1987-1996), and ICD 10 (from 1997), extracted by linking the MBR with the NPR using the unique personal identification number that each Swedish resident receives at birth or immigration. We followed the women from the index date until the AS diagnosis, death, or 31 December 2020, whichever occurred first.

Before 2001, only diagnoses in connection with hospitalization were recorded; after that, both diagnoses from hospital care and outpatient clinics were registered.

The MBR contains data on age, height, weight (registered at the first antenatal visit), smoking status (self-reported), and diabetes. Data on weight at the first antenatal visit have been registered since 1982 and were available in 70% of the women; data on height have been available since 1992 and were available in 80%. Between 1982 and 1989, weight >98 kg was recorded as 99 kg and, in these instances, weight was calculated by subtracting gestational weight gain from weight at delivery.

Data on comorbidities at baseline were obtained from the NPR (irrespective of position) and data about all-cause mortality were gathered from the National Cause of Death Register. The ICD codes used in the study are presented in Supplementary Table 1. Data on education were extracted from the longitudinal integration database for health insurance and labor market studies (LISA). In Sweden, data concerning employment and health insurance started in 1990, while in our study the follow-up time began in 1982, leading to a higher proportion of missing data in this variable.

BMI was calculated based on weight in kilograms divided by height in meters squared. We stratified the women into eight BMI categories, creating four categories in the group defined as normal weight by the WHO (<18.5 kg/m^2^, 18.5 kg/m^2^-<20 kg/m^2^, 20 kg/m^2^-<22.5 kg/m^2^, 22.5 kg/m^2^-<25 kg/m^2^), two categories in the group classified as overweight (25 kg/m^2^-<27.5 kg/m^2^, 27.5 kg/m^2^-<30 kg/m^2^) and two categories in the group defined as obesity (30 kg/m^2^-<35 kg/m^2^, ≥ 35 kg/m^2^; the latter category was defined as severe obesity).

### Statistical analysis

Baseline characteristics are presented as mean (standard deviation (SD)) or median (25^th^ percentile, 75^th^ percentile), as appropriate for continuous variables and frequency and percentage for categorical variables. We used Cox proportional hazards regression adjusted for age and baseline diabetes mellitus (type 1 and 2) to evaluate the risk of being diagnosed with AS in the eight BMI groups. The group with BMI 20 kg/m^2^-<22.5 kg/m^2^ was the reference category in calculating hazard ratios (HRs) and 95% confidence intervals (CIs). The assumption of proportional HRs was evaluated graphically using the scaled Schoenfeld residuals. Kaplan-Meier curves for time to AS diagnosis were created separately for each BMI group. A secondary analysis with and without AS of rheumatic or congenital etiologies (according to ICD codes in Supplementary Table 1) was conducted to confirm whether the results were unchanged regardless of etiology. The cumulative incidence function was used to estimate the marginal probability of death as competing event. The significance level was set at alpha <0.05.

The statistical analyses were performed using R version 4.2.2 ^26^ and SAS software version 9.4 (SAS Institute Inc, Cary, NC, USA)

## Results

After exclusions, 1,722,798 women were included, with a mean age of 28.3 ± 5.1 years, which was only marginally higher (28.8 ±5.4) in women with severe obesity (BMI≥35 kg/m^2^) (Table 1). The mean BMI in the whole cohort was 23.8 ± 4.2 kg/m^2^: 67.3% of the women had normal BMI (18.5-<25.0 kg/m^2^), 20.9% were overweight (BMI 25.0-30.0 kg/m^2^), 8.5% were obese, and 2.2% had severe obesity (BMI ≥35 kg/m^2^). Women in the lowest and highest BMI group had the highest percentage of smokers (25.1% and 26.8%, compared to 18.9% among those with a BMI of 20-22.5 kg/m^2^). In the whole cohort 31.7% had an education >12 years, higher in the BMI groups 22.5-<25 (33.8%) and 20-<22.5 kg/m^2^ (33.8%) and lower in the BMI groups <18.5 kg/m^2^ 23.3%) and ≥35 kg/m^2^ (25.6%).

**Table 1:** Study participants’ baseline characteristics by BMI group.

|  | BMI<18.50 | BMI 18.5-<20 | BMI 20-<22.5 | BMI 22.5-<25 | BMI 25-<27.5 | BMI 27.5-<30 | BMI 30-<35 | BMI ≥35 | Total |
| --- | --- | --- | --- | --- | --- | --- | --- | --- | --- |
|  | 58499 (3.4) | 171774 (10.0) | 538547 (31.3) | 448205 (26.0) | 236908 (13.8) | 122295 (7.1) | 108394 (6.3) | 38176 (2.2) | 1722798 |
| Age (years) Mean (SD) | 26.5 (5.0) | 27.5 (4.9) | 28.2 (4.9) | 28.5 (5.1) | 28.7 (5.2) | 28.7 (5.4) | 28.7 (5.4) | 28.8 (5.4) | 28.3 (5.1) |
| Median [Q1, Q3] | 26 (23,30) | 27 (24,31) | 28 (25,31) | 28 (25,32) | 28 (25,32) | 28 (25,32) | 28 (25,32) | 28 (25,32) | 28 (25,32) |
| Weight (kg) |  |  |  |  |  |  |  |  |  |
| **Mean (SD)** | 49.1 (4.1) | 53.8 (4.1) | 59.2 (4.8) | 65.5 (5.2) | 72.1 (5.9) | 78.8 (6.5) | 87.9 (7.6) | 105.3 (12.5) | 65.9 (12.3) |
| **BMI (kg/m2)** |  |  |  |  |  |  |  |  |  |
| **Mean (SD)** | 17.7 (0.7) | 19.4 (0.4) | 21.3 (0.7) | 23.7 (0.7) | 26.1 (0.7) | 28.6 (0.7) | 32.0 (1.4) | 38.4 (3.3) | 23.8 (4.2) |
| **Born in Sweden n (%)** | 42296 (72.3) | 136431 (79.4) | 441455 (82.0) | 362114 (80.8) | 185894 (78.5) | 93511 (76.5) | 84052 (77.5) | 30686 (80.4) | 1376439 (79.9) |
| **Deceased n (%)** | 1331 (2.28) | 3391 (1.97) | 9912 (1.84) | 8072 (1.80) | 4195 (1.77) | 2193 (1.79) | 2292 (2.11) | 686 (1.80) | 32072 (1.86) |
| **Smoking n (%)** | 14700 (25.1) | 36432 (21.2) | 101819 (18.9) | 85802 (19.1) | 49121 (20.7) | 27265 (22.3) | 26717 (24.6) | 10230 (26.8) | 352086 (20.4) |
| **Education** |  |  |  |  |  |  |  |  |  |
| **Missing** | 21534 (36.8) | 59719 (34.8) | 170068 (31.6) | 121650 (27.1) | 53819 (22.7) | 25415 (20.8) | 21401 (19.7) | 3949 (10.3) | 477555 (27.7) |
| **≤9 years** | 7129 (12.2) | 14043 (8.2) | 36988 (6.9) | 33785 (7.5) | 22272 (9.4) | 13847 (11.3) | 13842 (12.8) | 5911 (15.5) | 147817 (8.6) |
| **9-12 years** | 16194 (27.7) | 46587 (27.1) | 152788 (28.4) | 141418 (31.6) | 84532 (35.7) | 46895 (38.3) | 44275 (40.8) | 18541 (48.6) | 551230 (32.0) |
| **>12 years** | 13642 (23.3) | 51425 (29.9) | 178703 (33.2) | 151352 (33.8) | 76285 (32.2) | 36138 (29.5) | 28876 (26.6) | 9775 (25.6) | 546196 (31.7) |
| **MI, n (%)** | 1 (0.00) | 8 (0.00) | 18 (0.00) | 10 (0.00) | 14 (0.01) | 11 (0.01) | 6 (0.01) | 6 (0.02) | 74 (0.00) |
| **HF, n (%)** | 4 (0.01) | 15 (0.01) | 39 (0.01) | 47 (0.01) | 37 (0.02) | 18 (0.01) | 20 (0.02) | 13 (0.03) | 193 (0.01) |
| **AF, n (%)** | 14 (0.02) | 34 (0.02) | 119 (0.02) | 119 (0.03) | 61 (0.03) | 33 (0.03) | 36 (0.03) | 16 (0.04) | 432 (0.03) |
| **Cardiomyopathy, n (%)** | 9 (0.02) | 7 (0.00) | 31 (0.01) | 32 (0.01) | 23 (0.01) | 13 (0.01) | 14 (0.01) | 5 (0.01) | 134 (0.01) |
| **GUCH, n (%)** | 106 (0.18) | 261 (0.15) | 728 (0.14) | 676 (0.15) | 365 (0.15) | 192 (0.16) | 192 (0.18) | 69 (0.18) | 2589 (0.15) |
| **HT, n (%)** | 41 (0.07) | 141 (0.08) | 567 (0.11) | 599 (0.13) | 452 (0.19) | 308 (0.25) | 422 (0.39) | 241 (0.63) | 2771 (0.16) |
| **Stroke, n (%)** | 29 (0.05) | 115 (0.07) | 310 (0.06) | 290 (0.06) | 140 (0.06) | 100 (0.08) | 68 (0.06) | 30 (0.08) | 1082 (0.06) |
| **Ischemic stroke, n (%)** | 10 (0.02) | 44 (0.03) | 119 (0.02) | 108 (0.02) | 67 (0.03) | 39 (0.03) | 35 (0.03) | 19 (0.05) | 441 (0.03) |
| **Hemorrhagic stroke, n (%)** | 14 (0.02) | 51 (0.03) | 140 (0.03) | 136 (0.03) | 63 (0.03) | 42 (0.03) | 22 (0.02) | 9 (0.02) | 477 (0.03) |
| **PE, n (%)** | 30 (0.05) | 69 (0.04) | 320 (0.06) | 251 (0.06) | 226 (0.10) | 140 (0.11) | 142 (0.13) | 79 (0.21) | 1257 (0.07) |
| **DVT, n (%)** | 49 (0.08) | 151 (0.09) | 586 (0.11) | 563 (0.13) | 408 (0.17) | 262 (0.21) | 280 (0.26) | 138 (0.36) | 2437 (0.14) |
| **DM, n (%)** | 75 (0.13) | 285 (0.17) | 1670 (0.31) | 2322 (0.52) | 1693 (0.71) | 988 (0.81) | 955 (0.88) | 470 (1.23) | 8458 (0.49) |
| **RF, n (%)** | 30 (0.05) | 75 (0.04) | 251 (0.05) | 213 (0.05) | 94 (0.04) | 68 (0.06) | 66 (0.06) | 26 (0.07) | 823 (0.05) |
| **Cancer, n (%)** | 409 (0.70) | 1538 (0.90) | 5625 (1.04) | 5226 (1.17) | 2881 (1.22) | 1452 (1.19) | 1259 (1.16) | 417 (1.09) | 18807 (1.09) |

AF: atrial fibrillation, AS: aortic stenosis, BMI: body mass index, DM: diabetes mellitus, DVT: deep venous thrombosis,
GUCH: grown-up congenital heart diseases, HF- heart failure; HT: hypertension, IQR: interquartile range, MI: myocardial
infarction, PE: pulmonary embolism; RF: renal failure; SD: standard deviation

Baseline comorbidities were rare in this cohort of young women: most (except hemorrhagic stroke) had an increasing prevalence with higher BMI and the highest proportion of comorbidities among the obese. For example, hypertension was present in 0.63% of women with BMI ≥35 kg/m^2^ (compared to 0.11% in the reference group, BMI 20 -<22.5 kg/m^2^), diabetes in 1.23% compared to 0.31% and prior deep venous thrombosis in 0.36% compared to 0.11%. Cancer was less prevalent in the leanest BMI groups.

In the whole cohort, with a median follow-up of 19.5 years (minimum 0.08 – maximum 39.75), 2,513 women (0.15%) were diagnosed with AS, with the highest proportion among the obese and severely obese and the lowest in the group with BMI 18.5-20 kg/m^2^. The mean age of AS diagnosis decreased with higher BMI (from 50 years in the group with BMI 18.5-20 kg/m^2^ to 43 years in the severe obesity group). The time to diagnosis in the whole cohort was approximately 15-20 years, shorter for women with higher BMI (Figure 1). The relationship between AS and BMI was not linear (Figure 2), increasing dramatically at BMI >30 kg/m^2^.

**Figure 1:**
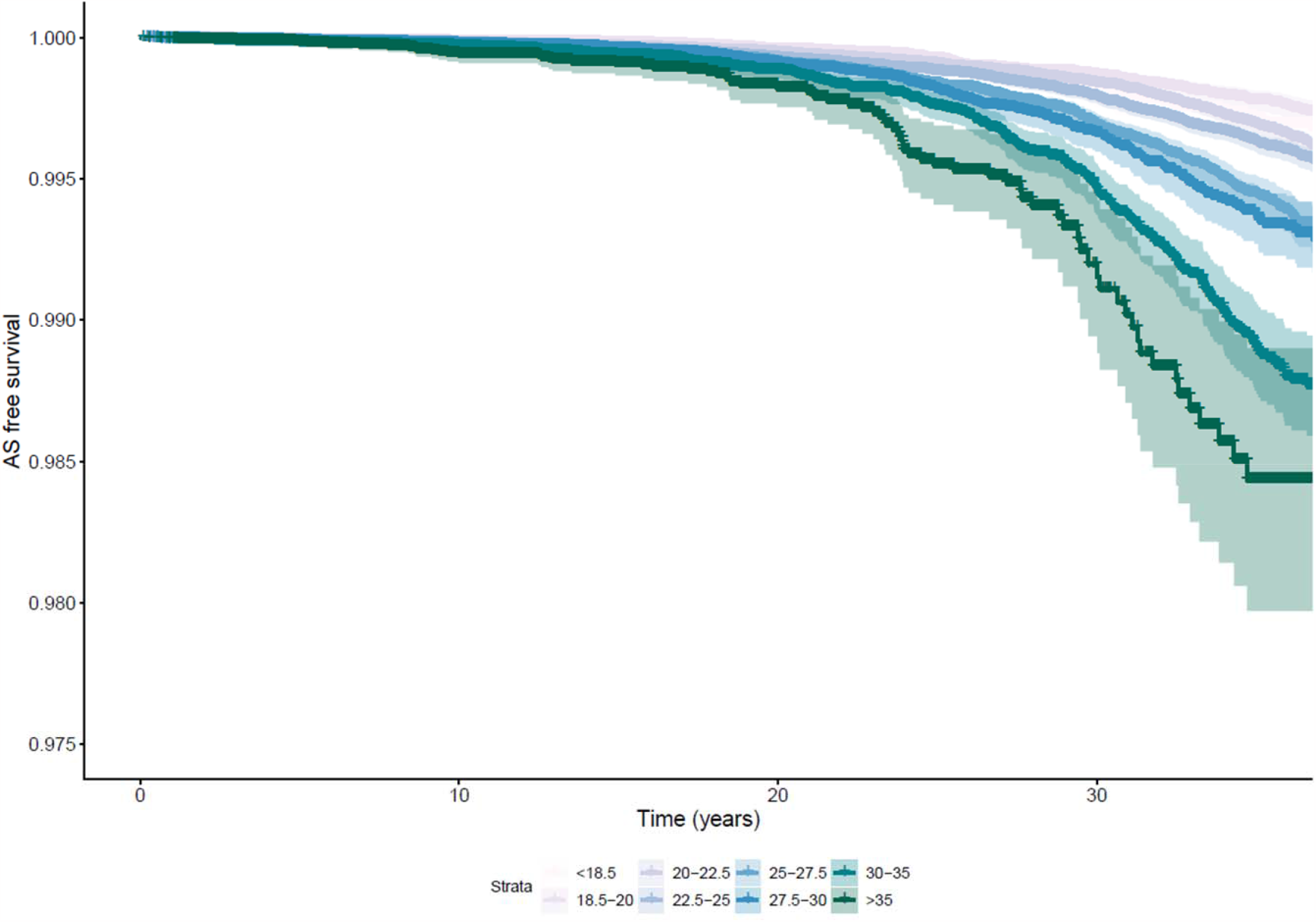
Kaplan-Meier curves for AS-free survival by BMI group.

**Figure 2:**
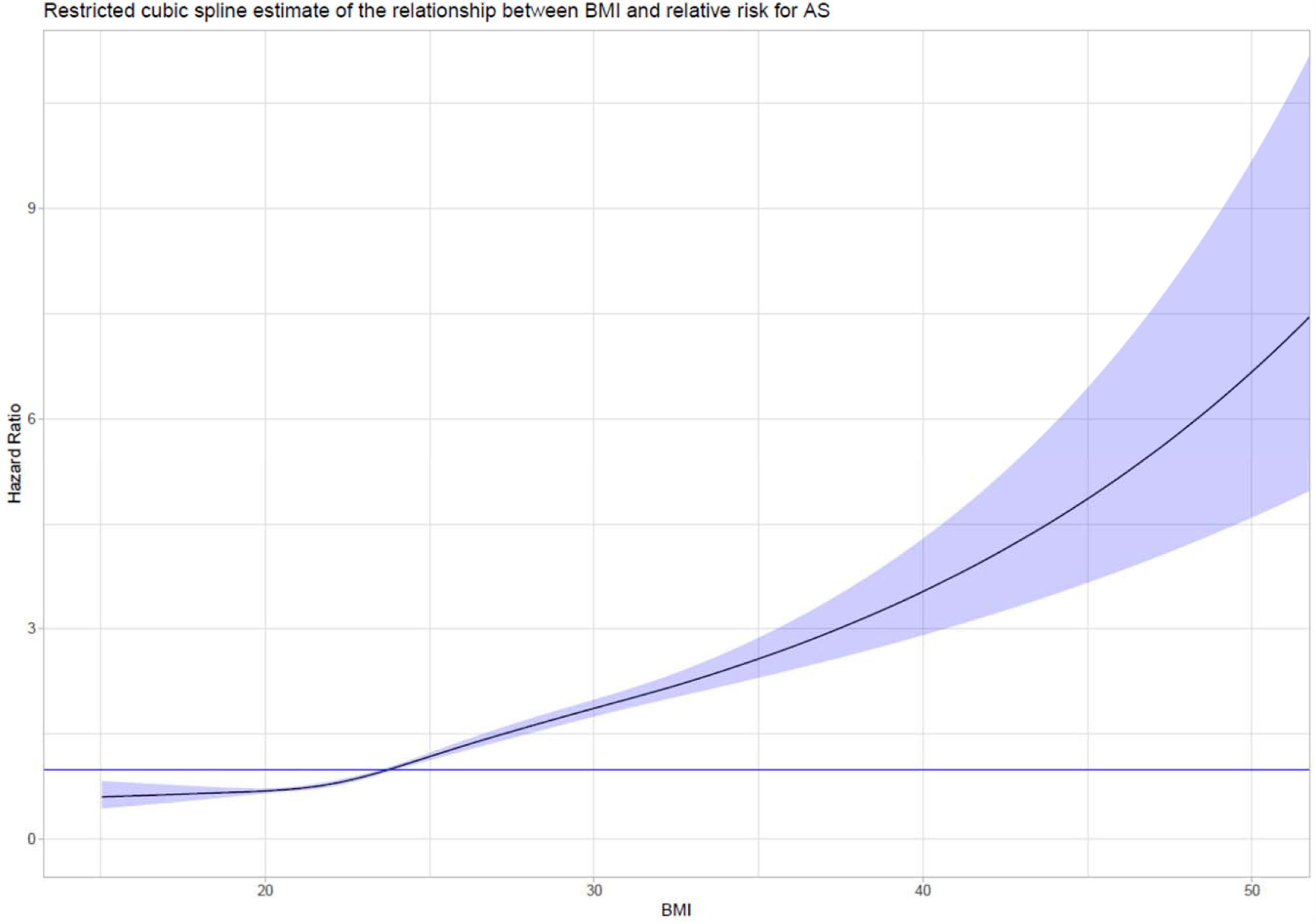
Restrictive cubic spline estimates of the relationship between BMI and relative risk for AS.

The risk of being diagnosed with AS during follow-up, adjusted for age and diabetes at baseline, with BMI 20 - <22.5 kg/m^2^ as reference, increased with higher BMI, from 1.17 (95% CI 1.05-1.31) times higher in those with BMI 22.5 -<25 kg/m^2^ to 2.78 (95%CI 2.41-3.21) times higher in women with obesity, with the highest HR 3.60 (95%CI 2.85-4.55) times higher in those with severe obesity (BMI group ≥35 kg/m^2^) (Figure 3). We also performed an analysis considering death as a competing risk and found that compared with women with BMI 20 -<22.5 kg/m^2^, women with BMI higher than 22.5 kg/m^2^ had increased risk to be diagnosed with AS, the highest risk being in obese women (HR 3.49, 95%CI 2.62-4.65).

**Figure 3:**
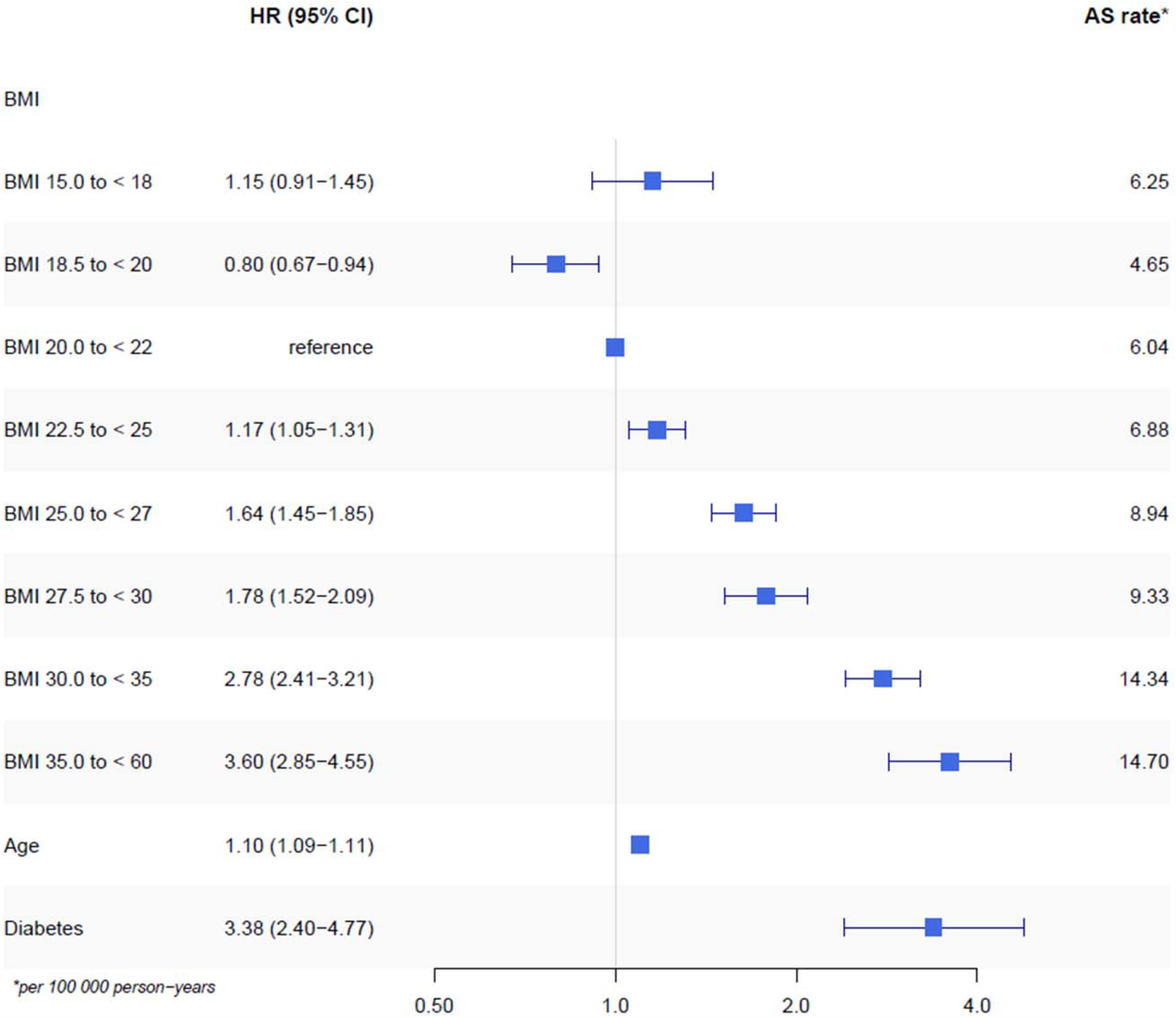
Forest plot of the risk for AS adjusted for age and DM according to BMI group:

In a secondary analysis, in which we excluded women with rheumatic or congenital etiology (n=162), we obtained essentially the same results as for AS of degenerative etiology, with a higher risk of AS in those with higher BMI.

## Discussion

In this study we aimed to investigate whether the association of high BMI with AS that we found in two previous studies in men also applies to young women. We found that even women in the high-normal BMI group (22.5 -<25 kg/m^2^) had a higher risk of being diagnosed with AS, which increased with higher BMI, reaching a risk of over three times higher in women with severe obesity. This association between AS and BMI, even in individuals with what is considered a normal BMI (22.5-<25 kg/m^2^), was also found in one of our previous studies in men ^13^. We also noted that women with higher BMI are diagnosed with AS at a younger age, likely contributing, albeit to a comparatively minor extent, to increased morbidity and mortality in overweight or obese people.

Studies on the association between BMI and AS have not been consistent. Some studies found a positive correlation between BMI and the risk for AS. For example, a French study of patients with severe AS (mean age 69 years) found that these patients had significantly higher BMI than controls ^20^. A more recent study that used ICD codes to identify patients with AS reached the same conclusion in men (mean age at baseline 59 years) and women (mean age at baseline 60 years) ^27^. A recent Danish study using Mendelian randomization, showed that genetically increased BMI was causally associated with an increased risk of AS, further strengthening causal inference ^21^. Our study confirms these findings in a younger cohort, including in the high-normal BMI group (22.5-<25 kg/m^2^).

However, some studies did not find any association between obesity and AS. For instance, a sub-study of the SEAS study (Simvastatin and Ezetimibe in Aortic Stenosis), a randomized controlled trial of Simvastatin and Ezetimibe versus placebo in patients with initially asymptomatic AS ^22^, did not find a correlation between obesity or overweight with AS. However, because individuals with diabetes mellitus or hypercholesterolemia were excluded, a substantial proportion of patients with obesity were likely not included. Another study in which no association between AS and obesity was found included individuals enrolled in the Cardiovascular Health Study. However, this study was cross-sectional, lacking the possibility to ascertain a time-dependent correlation between obesity, a risk factor that acts insidiously, and a slowly progressive disease such as AS ^23^. In contrast to our findings, one study concluded that older adults with low BMI could have a higher risk of aortic valve calcification ^24^. This study, however, was also cross-sectional, and the findings not reproduced in other studies.

The pathophysiological process that leads to AS begins with a lesion on the valves caused by a disturbance in the hemodynamic environment, followed by lipid infiltration, inflammation, and calcification of the aortic valves ^28^. Also, persons with obesity have a volume overload and a higher prevalence of hypertension ^29,30^, resulting in higher turbulence around the aorta valve and the possibility of endothelial lesions. In addition, obesity induces low-grade inflammation, leading to more available inflammatory cells to infiltrate the subendothelial lesion ^31^. Lipid metabolism is also disturbed in people with high BMI ^29^, and lipoproteins play an important role in incipient aortic lesions ^28,32^.

The cumulative incidence of AS in our study was low due to the young age of the women in the study, whereas AS is usually diagnosed in older people^1^, and is, additionally, more prevalent in men ^33^. Considering that congenital and rheumatic AS have different pathophysiologies, we repeated the analyses by excluding AS of rheumatic and congenital etiology, obtaining the same results. Thus, the risk of developing AS increases in those with higher BMI, regardless of the inclusion of AS from other than degenerative etiology.

Although it seems that obesity can elevate the risk of AS, it also acts as a protective factor in patients already diagnosed with AS. As an example, in patients undergoing TAVR (transcatheter aortic valve replacement), obese patients have lower short- and long-term overall mortality after TAVR than normal-weight patients ^34^. In addition, another study showed that obesity after TAVR does not render a higher risk of post-operative mortality compared to patients with normal BMI and that patients with BMI <20 kg/m^2^ had the highest post-operative mortality ^35^. However, a lower BMI is likely a marker of frailty and poor prognosis in older patients with severe AS.

With longer life expectancy, the proportion of older adults worldwide is estimated to increase, mostly in the age groups 70-79 years and >80 ^4^, the age groups with the highest prevalence of AS ^1^. Therefore, severe AS is expected to increase three-fold until 2060 ^4^. In addition, obesity is also increasing in both developed and undeveloped countries ^9^. Consequently, if other studies confirm the association of obesity with AS, the effort to prevent this serious cardiovascular disorder becomes even more critical.

Because AS is a condition that progresses slowly, with death a significant competing risk, it might be difficult to show to which extent different approaches could delay or prevent AS. Even more difficult would be to demonstrate that maintaining a normal or low-normal weight might prevent AS or that weight-reducing measures in patients with higher BMI could lead to fewer AS cases. However, in this era, with an increased proportion of individuals with early overweight and obesity, considering that there is no medical treatment that can ameliorate AS prognosis, prevention by maintaining a low-normal or normal BMI during life is likely becoming even more important.

### Strengths and limitations

The major strength of this study was the inclusion of a large number of younger women, a subpopulation that has rarely been examined. We also verified our results excluding cases of specific non-degenerative etiologies and also by including death as a competing risk.

However, our study has limitations that should be acknowledged. First, by including only women, the external validity of our findings is limited. However, this study adds important information to previous studies, some of which were only conducted in men. Second, only women who gave birth were included. However, to our knowledge there are no relevant differences between women who give birth and those who do not concerning the risk of acquiring AS. Third, the study has the inherent limitations of a register study, where the AS cases were identified using ICD codes and no echocardiographic data or data on AS severity were available. Fourth, a measuring bias for BMI might be present because in some periods during the follow-up weight or height was self-reported, and between 1982 and 1989, all weights >98 kg were coded as 99 kg. However, the weights during this period were adjusted.

## Conclusion

The risk of developing AS in women increases in those with higher BMI, even in the high-normal group, regardless of etiology. These findings underline the importance of maintaining a normal BMI (<25 kg/m^2^) and even a BMI <22.5 kg/m^2^.

## Data Availability

The data that support the findings of this study are available on request from the corresponding author, SK.

## Abbreviations

AS: aortic stenosis
BMI: body mass index
CI: confidence interval
HR: hazard ratio
ICD: International Classification of Diseases
LISA: Longitudinal integration database for health insurance and labor market studies
MBR: Swedish Medical Birth Register
NPR: Swedish National Patient Register
SD: standard deviation
SEAS: Simvastatin and Ezetimibe in Aortic Stenosis
TAVR: transcatheter aortic valve replacement

## Author contribution

SK, AR, TZ, MF, and ML conceived the project; SK drafted the manuscript; SK and TZ performed statistical analysis; AR and ML were responsible for funding acquisition; AR, ET, TS, and CB collected the data; SK, TZ, AR, ET, ML, MF, CB, and TS critically revised the manuscript. All authors gave final approval and agreed to be accountable for all aspects of the work, ensuring integrity and accuracy.

## Acknowledgements and Funding

This work was supported by grants from the Swedish state under an agreement between the Swedish government and the county councils concerning economic support of research and education of doctors [ALFGBG-966211]; [ALFGBG-971608], the Swedish Heart and Lung Foundation [2021-0345], and the Swedish Research Council [2018-02527; VRREG 2019-00193].

## Figures and tables legends

Supplementary Figure 1: Flow chart of the study participants

Supplementary Table 1: ICD codes used in the study

